# A city-level analysis of air pollution, climate and COVID-19 early spread during the Spanish lockdown

**DOI:** 10.1101/2020.08.09.20171041

**Authors:** Álvaro Briz-Redón, Carolina Belenguer-Sapiña, Ángel Serrano-Aroca

**Author notes:** These authors contributed equally (Álvaro Briz-Redón, Carolina Belenguer-Sapiña).

## Abstract

The COVID-19 outbreak has escalated into the worse pandemic of the present century. The fast spread of the new SARS-CoV-2 coronavirus has caused devastating health and economic crises all over the world, with Spain being one of the worst affected countries in terms of confirmed COVID-19 cases and deaths per inhabitant. In this situation, the Spanish Government declared the lockdown of the country. The variations of air pollution in terms of fine particulate matter (PM_2.5_) levels in seven cities of Spain are analyzed here considering the effect of meteorology during the national lockdown. The possible associations of PM_2.5_ pollution and climate with COVID-19 accumulated cases were also analyzed. While the epidemic curve was flattened, the results of the analysis show that the 4-week Spanish lockdown significantly reduced the PM_2.5_ levels in only one of the cities despite the drastically reduced human activity in good agreement with our previous study of changes in air quality in terms of CO, SO_2_, PM_10_, O_3_ and NO_2_ levels. Furthermore, no associations between either PM_2.5_ exposure or environmental conditions and COVID-19 transmission were found during the early spread of the pandemic.

## 1. Introduction

The global pandemic of the SARS-CoV-2 coronavirus causing COVID-19, first announced in Wuhan, China, in December 2019(WHO, 2020a), continues to spread in more than 200 countries(WHO, 2020b). Spain confirmed 314,362 COVID-19 infections and 28,503 death people(ISCIII, n.d.) on August 9, 2020, which is one of the largest mortality rates of this emerging disease. Due to the growing pandemic, governments have been forced to react with lighter or stronger control measures influenced by their cultural backgrounds(Chiu et al., 2020). The government of Spain declared a lockdown on March 14, 2020. Extraordinary control policies were executed in an effort to reduce COVID-19 transmission. At the beginning of the lockdown, from March 15 to March 29, 2020, named here as *minor lockdown*, people were recommended to work from home, and other measures such as travel restrictions, isolation, and quarantine of patients, cancellation of private and public events, online education, the closing of restaurants, bars and pubs, or the prohibition of public congregations, were imposed. During those two weeks, only essential products could be sold in supermarkets and drugstores, and only a small amount of activities were allowed. During the last two weeks of the lockdown, from March 30 to April 12, 2020, named here as *major lockdown*, the Spanish Government imposed more severe restrictions due to the national emergency caused by the collapse of the health system. This situation forced people to stay at home (except for very limited purposes) and only essential work such as healthcare and social care sectors, police and armed forces, water and electricity supply was allowed. Besides, important industrial activities such as construction were forbidden. These unprecedented measures gave positive results and flattened the epidemic curve after a month of lockdown(Tobías, 2020). In short, the pandemic has definitely caused significant behavioral variations in society and could provide a useful insight into how we can make our production methods more sustainable(Sarkis et al., 2020). The impact of particulate matter (PM) on health is well-known(Manisalidis et al., 2020) in terms of its effects on morbidity and mortality. Two sizes of particulate matter are used to analyze air quality; fine particulate matter or PM_2.5_, with a diameter of 2.5 μm or less, and coarse particles or PM_10_, with a diameter of 10 μm or less. However, the former is more worrying because their small size allows them to penetrate deeper into the human respiratory system via inhalation, which can potentially promote respiratory diseases(Chai et al., 2019; Dominici et al., 2006; Horne et al., 2018) such as COVID-19. Thus, it is of note that citizens exposed to a high concentration of PM_2.5_ are more prone to developing chronic respiratory diseases favorable to infective agents(Mehmood et al., 2020). Long-term exposure to these small particles can produce a chronic inflammatory stimulus, especially in unhealthy people(Conticini et al., 2020).In addition, short-term exposure to PM_2.5_ particles may also increase susceptibility to infections(Chen et al., 2020). Indeed, this type of pollution can harm human airways, promoting viral infections, and diminish the immune response(Liu et al., 2019; Mehmood et al., 2020; Sedlmaier et al., 2009). The World Health Organization’s air quality guidelines recommend that PM_2.5_ concentration should not exceed a 10 μg/m^3^ annual mean and 25 μg/m^3^ 24-hour mean(World Health Organization, 2018). Due to combustion processes, road traffic is the main emission source of atmospheric PM_2.5_(Nicolás et al., 2020). Other sources of PM_2.5_ in urban environments include Sahara dust events(Fenech and Aquilina, 2020; Nicolás et al., 2020), shipping(Viana et al., 2020), secondary inorganic aerosol or biomass burning(Liu et al., 2020), combustion processes in thermal power stations and other industrial sectors, the transport of anthropogenic aerosols from central Europe to Mediterranean areas, and certain agricultural activities(García et al., 2019). Spain, which still consumes a considerable amount of fossil fuels, is quite near to the Sahara Desert and has approximately 47 million inhabitants. Thus, it is in a high-risk zone of pollution episodes(Lemou et al., 2020). The Mediterranean zone has been identified as a crossroads of air masses with many kinds of aerosols caused by many anthropogenic and natural sources, such as the resuspension of dust from Africa, the production of sea salt, industrial and urban aerosols, fires, and smoke from Eastern Europe(Lemou et al., 2020). The zone is also known for its intensive shipping traffic, near a densely populated and environmentally sensitive area with low atmospheric dispersion(Viana et al., 2020). Since Spain has been positioned as one of the ten most climatically differing nations (Ministerio de Medio Ambiente y Medio Rural y Marino. AEMET, 2011), it is necessary to take meteorological factors(Briz-Redón and Serrano-Aroca, 2020a) into account when evaluating atmospheric pollutant concentrations. We have recently demonstrated that the Spanish lockdown did not decrease air pollution (NO_2_, CO, SO_2_, and PM_10_) considering meteorological factors on the pollutants’ levels(Briz-Redón et al., 2020). Furthermore, the O3 levels were generally increased during this period. In the current paper, we focus on the variations of air pollution in terms of fine particulate matter, wich could be associated with the COVID-19 spread(Mehmood et al., 2020), in several representative Spanish cities during the lockdown using PM_2.5_ pollution and COVID-19 data. Meteorological parameters, which significantly affect air pollution (Shi et al., 2018; M. Wang et al., 2020; Wang et al., 2019) have also been taken into account. Several studies in the same research line have shown evidence of significant reductions of severe PM_2.5_ pollution during COVID-19 lockdowns in other countries, such as India(Mahrati et al., 2020; Sharma et al., 2020) and Malaysia(Abdullah et al., 2020). However, it has been reported an increased of the PM_2.5_ levels in Tehran (Faridi et al., 2020) due to the reduction of public transportation use and thus increase of traveling by private vehicle, and in other countries such as China, which probably applied one of the strictest lockdown measures, the fine particulate matter in the atmosphere was not significantly reduced(P. Wang et al., 2020). On the other hand, COVID-19 daily new cases have been positively related with PM_2.5_ levels in 355 municipalities in the Netherlands(Cole et al., 2020), in Milan(Zoran et al., 2020) and in China(Zhu et al., 2020). However, additional research is needed to test the exploratory associations found between air pollution and COVID-19 prevalence so far(Hendryx and Luo, 2020). Therefore, in this paper, we also explore the possible impact of PM_2.5_ exposure and the effect of environmental conditions (temperature, wind, precipitation, sunlight, pressure) on the COVID-19 early spread.

## 2. Data

### 2.1. PM_2.5_ pollution

Seven major Spanish cities were considered in this study (see Figure 1).Longitude

**Figure 1.**
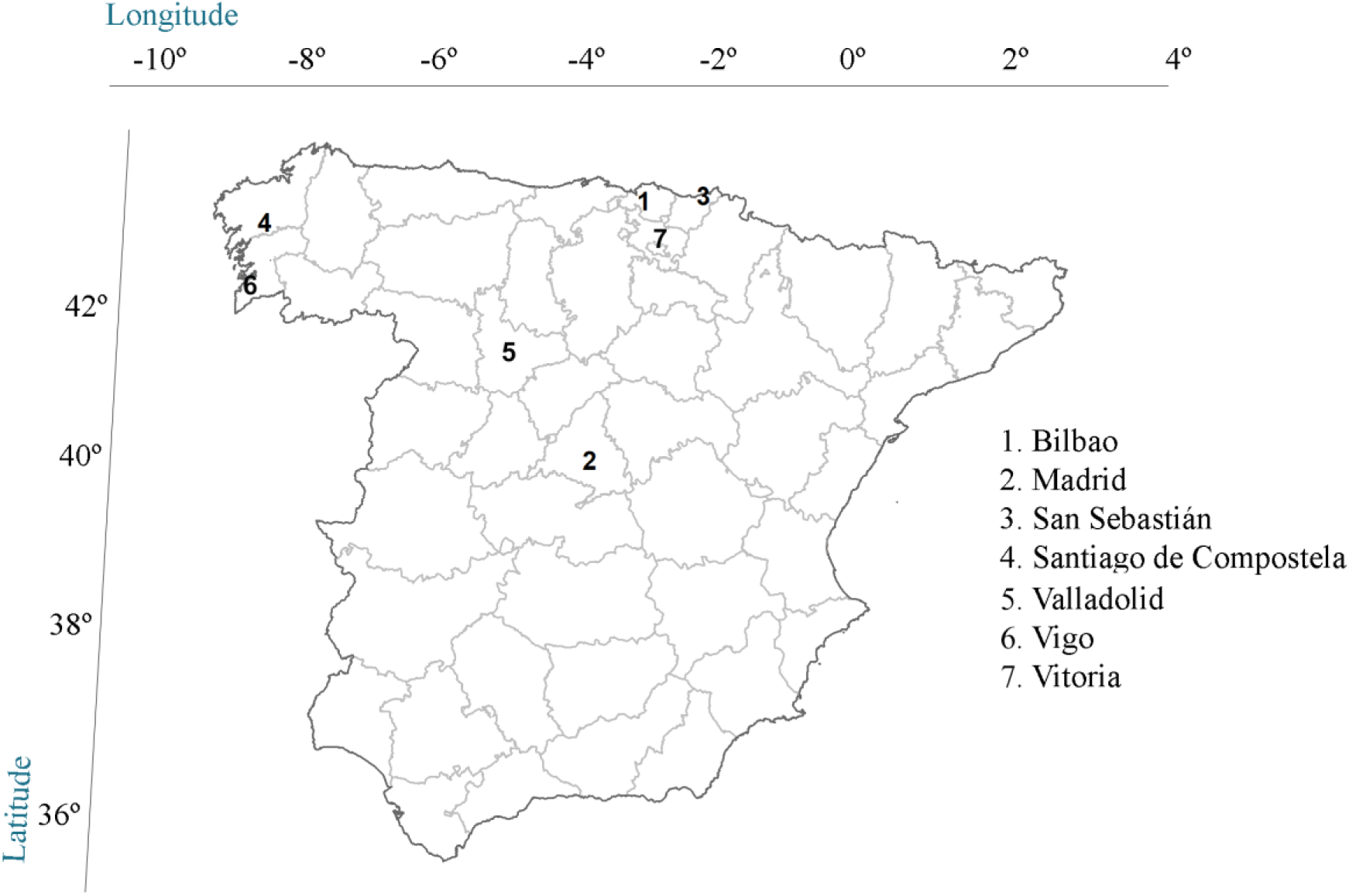
Map of Spain with the seven cities considered for the PM_2.5_ pollution analysis. Location coordinates are those of the pollution station. The provincial borders are shown in grey.

Table 1 shows that the information of each selected city with populations (on January 1, 2019) that vary from 97,260 to 3,266,126 inhabitants(INE, 2019).

**Table 1.**
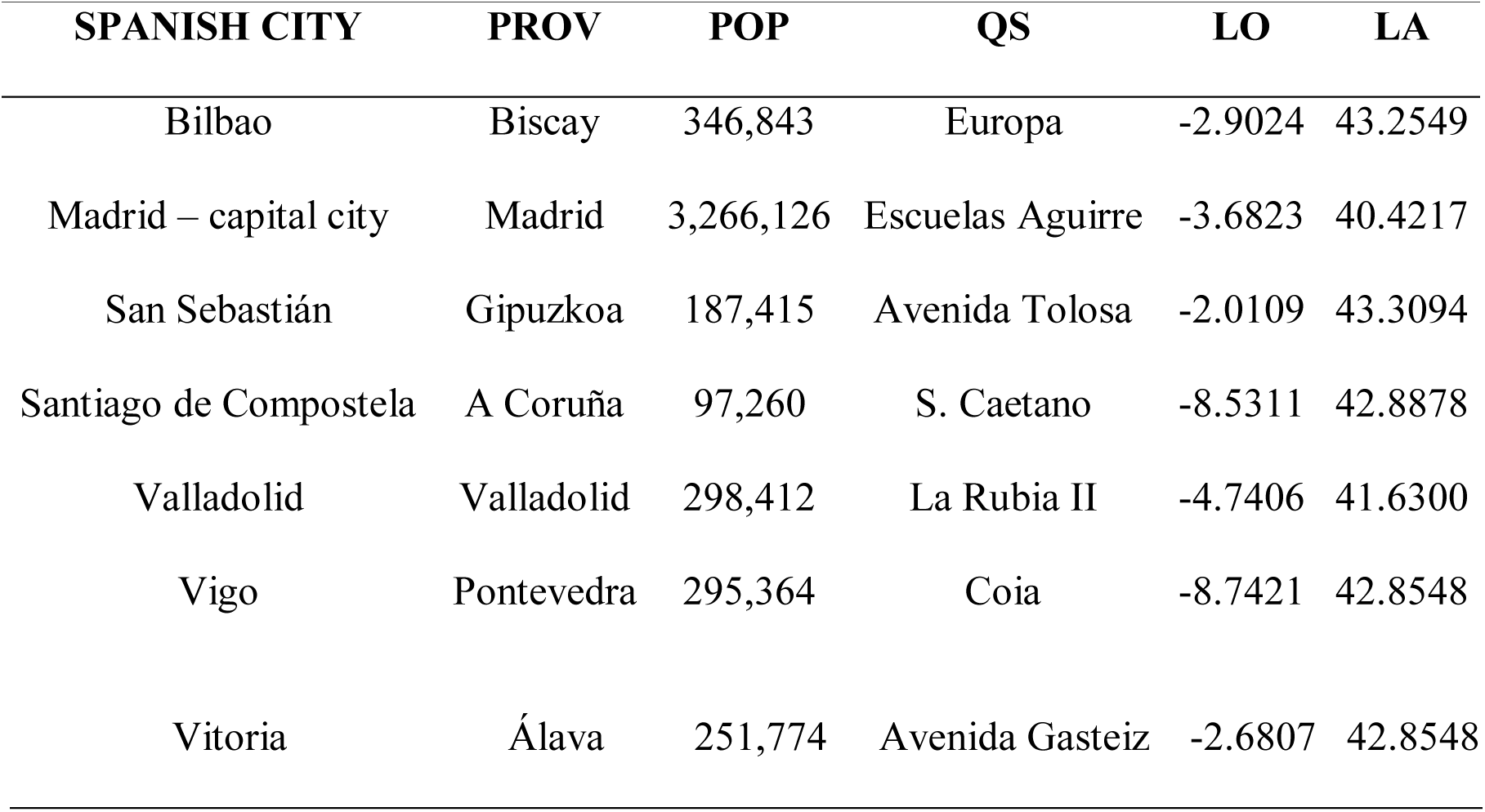
Spanish city, province (PROV), population (POP, on January 1, 2019), longitude (LO) and latitude (LA) of the air quality stations (QS) selected for the study.

Table 2 provides a city-level statistical summary of the PM_2.5_ levels in each of the periods considered for the analysis.

**Table 2.** City-level statistical summary of PM_2.5_ levels in terms of mean value, 1^st^ quartile (1^st^ Q), and 3^rd^ quartile (3^rd^ Q) at each of the different periods (normal, minor and major lockdown) considered for the analysis

| Spanish city | Normal period |  |  | Minor lockdown |  |  | Major lockdown |  |  |
| --- | --- | --- | --- | --- | --- | --- | --- | --- | --- |
|  | Mean | 1<sup>st</sup> Q | 3<sup>rd</sup> Q | Mean | 1<sup>st</sup> Q | 3<sup>rd</sup> Q | Mean | 1<sup>st</sup> Q | 3<sup>rd</sup> Q |
| Bilbao | 9.13 | 6.00 | 12.50 | 13.93 | 11.00 | 17.00 | 8.14 | 6.25 | 9.75 |
| Madrid | 11.51 | 6.00 | 14.00 | 6.60 | 4.50 | 8.50 | 7.00 | 5.00 | 9.00 |
| San Sebastián | 8.95 | 6.00 | 11.00 | 13.17 | 10.75 | 15.50 | 8.64 | 7.00 | 10.00 |
| Santiago de Compostela | 12.64 | 8.20 | 17.00 | 15.18 | 13.00 | 18.00 | 9.01 | 7.23 | 9.85 |
| Valladolid | 11.30 | 6.50 | 15.00 | 11.40 | 8.50 | 13.00 | 6.14 | 5.00 | 7.00 |
| Vigo | 9.11 | 6.45 | 12.00 | 11.93 | 9.20 | 13.50 | 6.21 | 5.00 | 6.78 |
| Vitoria | 7.33 | 4.25 | 10.00 | 10.87 | 9.00 | 13.50 | 6.36 | 4.50 | 7.00 |

PM_2.5_ pollution data was obtained from official web pages: Bilbao(Euskadi.eus, 2020); Madrid(Ayuntamiento de Madrid, 2020); San Sebastián(Euskadi.eus, 2020); Santiago (Xunta de Galicia, 2020); Valladolid(Junta de Castilla y León, 2020); Vigo(Xunta de Galicia, 2020) and Vitoria(Euskadi.eus, 2020). This pollution data was obtained from a traffic station of each city (see Table 1), whose pollution levels are mainly the consequence of traffic emissions(UK Dep. for Environment Food and Rural Affairs, 2020). Every day, PM_2.5_ (in μg/m^3^) were collected from each station from March 4 to April 14, 2019, and from March 2 to April 12, 2020, using the sampling method defined by the current Directive 2015/1480(2015/1480/UE, 2015) instead of the previous(2008/50/CE, 2008), following the gravimetric method of determining PM_2.5_ mass fraction in suspended particulate matter (EN 12341:2014, 2014). The measurements are commonly performed with active samplers operating at 2.3 m^3^/h over a sampling period of 24 h. The range of application of this European Standard is from 1 μg/m^3^ (detection method limit) up to approximately 120 μg/m^3^.

### 2.2. Meteorological data

The API of the OpenData platform of the State Meteorological Agency was used to download the meteorological data necessary for this analysis. One meteorological station per city was selected for the meteorological variables considered in this study: temperature, precipitation, wind velocity, min, and max atmospheric pressure, and sunlight time, that is, number of hours with a solar irradiance over 120W/m^2^. The minimum pressure was discarded for the analysis because of the high correlation (very close to 1) between the maximum and minimum pressure values. Table 3 provides a statistical summary of the meteorological variables considered.

**Table 3.** Summary of the meteorological variables (temperature (T), Precipitation (PR), Wind speed (WS), sunlight hours (SH) and maximum pressure (MP)) considered for the study in terms of mean value, 1^st^ quartile (1^st^ Q), and 3 ^rd^ quartile (3 ^rd^ Q) during the entire study period.

| Spanish city | T (°C) |  |  | PR(mm) |  |  | WS (km/h) |  |  | SH (h) |  |  | MP (hPa) |  |  |
| --- | --- | --- | --- | --- | --- | --- | --- | --- | --- | --- | --- | --- | --- | --- | --- |
|  | Mean | 1<sup>st</sup>Q | 3<sup>rd</sup>Q | Mean | 1<sup>st</sup>Q | 3<sup>rd</sup>Q | Mean | 1<sup>st</sup>Q | 3<sup>rd</sup>Q | Mean | 1<sup>st</sup>Q | 3<sup>rd</sup>Q | Mean | 1<sup>st</sup>Q | 3<sup>rd</sup>Q |
| Bilbao | 12.2 | 10.2 | 14.2 | 3.1 | 0.0 | 3.1 | 3.5 | 2.2 | 3.9 | 4.5 | 0.9 | 7.6 | 1017.1 | 1012.6 | 1022.7 |
| Madrid | 11.3 | 9.7 | 13.2 | 1.4 | 0.0 | 0.5 | 3.3 | 1.9 | 4.2 | 7.1 | 3.4 | 11.0 | 952.8 | 949.2 | 957.8 |
| San Sebastián | 11.2 | 9.3 | 13.0 | 3.92 | 0.0 | 5.1 | 4.5 | 2.8 | 5.3 | 5.3 | 1.4 | 9.0 | 990.7 | 986.3 | 996.2 |
| Santiago de Compostela | 10.3 | 8.4 | 12.5 | 4.2 | 0.00 | 3.4 | 2.9 | 1.9 | 3.6 | 5.9 | 2.4 | 10.4 | 978.5 | 974.2 | 983.3 |
| Valladolid | 10.9 | 7.9 | 11.6 | 1.2 | 0.0 | 1.2 | 2.2 | 1.4 | 2.8 | 7.3 | 4.3 | 10.6 | 935.9 | 932.0 | 941.1 |
| Vigo | 11.9 | 10.0 | 13.5 | 5.2 | 0.0 | 3.9 | 3.5 | 2.8 | 3.9 | 6.8 | 2.7 | 10.6 | 990.9 | 986.3 | 995.4 |
| Vitoria | 8.6 | 6.6 | 10.4 | 1.4 | 0.0 | 1.0 | 3.6 | 2.5 | 4.2 | 6.2 | 3.0 | 9.8 | 961.7 | 957.6 | 966.9 |

### 2.3. COVID-19 data

COVID-19 data was also downloaded from multiple local and regional webpages: Bilbao(Euskadi, 2020), Madrid(Datos Abiertos Comunidad de Madrid, 2020), San Sebastián (Euskadi, 2020), Santiago de Compostela(Galicia, 2020), Valladolid(JCyL, 2020), Vigo(Galicia, 2020), and Vitoria(Euskadi, 2020). Data was collected for the period comprised from March 18 to April 12, 2020, and corresponded to the number of accumulated COVID-19 cases in each city.

## 3. Methods

### 3.1. R programming language

The R programming language(Team, 2020) was used for the statistical analysis with several R packages: *effects*(Fox and Weisberg, 2018), *ggplot2*(Wickham, 2016), *INLA* (Lindgren and Rue, 2015; Rue et al., 2009), *lubridate*(Grolemund and Wickham, 2011), *RCurl*(Lang and the CRAN team, 2015), sjPlot(Ludecke, 2016) and *XML*(Lang, 2020).

### 3.2. Modeling pollution levels

In order to discriminate between the effects of meteorology and lockdown, the meteorological variables (temperature, precipitation, wind velocity, sunlight time, or atmospheric pressure), which modify pollutant levels (Venter et al., 2020), were considered in the statistical model. Weekend days were also considered because of their reduced pollutant levels as a result of less road traffic. Thus, the daily PM_2.5_ pollutant levels of the seven cities were fitted through a statistical model considering meteorological variables, weekend days, and lockdown periods. The PM_2.5_ pollutant levels for a city *i* on date *t* were modeled by Equation (1), including quadratic terms capable of capturing non-linear relationships between the meteorological variables and the PM_2.5_ level.

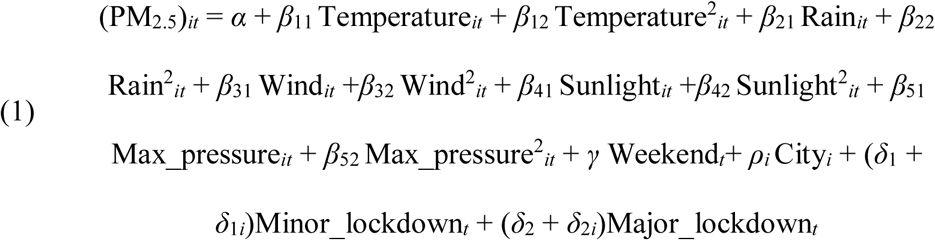

In this regression model, *α* is the global intercept of the model, *β_kj_ (k* = 1, 2, 3, 4, 5 and *j* = 1, 2) quantifies the corresponding meteorological covariate effect (in quadratic form, or not) on the (PM_2.5_)*_it_* values, *γ* quantifies weekend days effect on the (PM_2.5_)*_it_* values, *ρ_i_* represents the city-specific effect at no lockdown PM_2.5_ levels, *δ*_1_ and *δ*_2_ indicate the overall *lockdown* effect *(minor* and *major*, respectively) on (PM_2.5_)*_it_* levels, and *δ*_1_*_i_* and *δ*_2_*_i_* indicate the city-specific effect *(minor* and *major*, respectively) on the (PM_2.5_)*_it_* level. Bank holidays were computed as weekend days. Therefore, this model estimates the variations of PM_2.5_ pollution levels because of lockdown while simultaneously accounting for week-day and meteorological effects.

### 3.3. Modeling COVID-19 spread

The association between COVID-19 spread and the PM_2.5_ levels and environment was also studied for all the cities included in the study. Hence, the number of accumulated COVID-19 cases in each city was modelled in terms of each of the environmental covariates available (temperature, rain, wind velocity, sunlight time, maximum atmospheric pressure, and PM_2.5_ level) through a Poisson model. Thus, the accumulated COVID-19 cases on date *t*, *y_t_*, was modelled as follows:

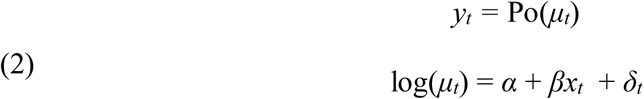

where *α* is the global intercept of the model, *x_t_* represents the environmental covariate, *β* refers to the coefficient that measures the magnitude of the effect of *x_t_* on log(*μ_t_*), and *δ_t_* is a temporally-structured effect for day *t* to control for serial correlation, which was defined by a first-order random walk. Lagged covariate effects were considered in the model by replacing the term *x_t_* by *x_t-7_*, and *x_t-14_*, which allows assessing the possible effect of the environmental conditions on the previous two weeks on the total number of cases reported on date *t*. The model described by Equation (2) was fitted using the Integrated Nested Laplace Approximation (INLA) method(Lindgren and Rue, 2015; Rue et al., 2009).

## 4. Results and discussion

### 4.1. Global effects

A stepwise algorithm was applied to the regression model represented by Equation (1) to find the subset of the variables providing the best model in terms of the Akaike information criterion. The overall variables included in this model and the coefficients associated with each one are shown in Table 4.

**Table 4.** Global effects estimated between variables and fine particulate matter with Equation (1). The estimated model coefficients are shown together with the associated *p*-values.

|  | **Estimate** | **p-value** |
| --- | --- | --- |
| **(Intercept) ($\alpha$)** | -943.8003 | 0.0521 |
| **Temperature ($\beta_{11}$)** | 0.7071 | 0.0393 |
| **Temperature<sup>2</sup> ($\beta_{12}$)** | -0.0241 | 0.1202 |
| **Wind speed ($\beta_{31}$)** | -1.0710 | 0.0002 |
| **Wind speed<sup>2</sup> ($\beta_{32}$)** | 0.0402 | 0.1156 |
| **Sunlight hours<sup>2</sup> ($\beta_{42}$)** | 0.0322 | 0.0000 |
| **Max Pressure ($\beta_{51}$)** | 1.8652 | 0.0608 |
| **Max Pressure<sup>2</sup> ($\beta_{52}$)** | -0.0009 | 0.0717 |
| **Weekend day ($\gamma$)** | 0.8308 | 0.0215 |
| **Minor lockdown day ($\delta_1$)** | 4.3791 | 0.0001 |
| **Major lockdown day ($\delta_2$)** | -1.4703 | 0.2328 |

First, it is worth noting that the two precipitation-related variables were discarded, suggesting the absence of an association between precipitation and PM_2.5_ values. The model also suggests that temperature, wind speed, the number of sunlight hours, and the maximum pressure have a quadratic relationship with PM_2.5_ levels. Thus, for the range of values attained by these meteorological variables during the period under study, the coefficients estimated for the linear and quadratic forms of the variables indicate that higher values of either temperature, sunlight hours, or maximum pressure are associated with higher PM_2.5_ levels, whereas higher wind speed values are associated with lower PM_2.5_ levels. The results obtained mostly agree with other studies in the literature. Thus, regarding the positive association of PM_2.5_ with temperature and sunlight hours, dry sunny weather frequently leads to prevent the vertical dispersion of pollutants due to thermal inversion(Mao et al., 2020) and increases their concentration, generating significant smog episodes. Indeed, sunny weather also favors photochemical reactions(P. Wang et al., 2020; Xu et al., 2020) while dry conditions prolong aerosols and atmospheric loadings(Lemou et al., 2020). The increase of wind speed can also decrease PM_2.5_ levels(Radzka, 2020; Yousefian et al., 2020), while lower wind speeds can promote a reduction of particulate matter in the air because of a significant increase of deposition(Xu et al., 2020). The rise of atmospheric pressure seems to positively affect PM formation(Hoque et al., 2020).

### 4.2. Coefficients of determination

Table 5 shows the coefficients of determination (*R*^2^) for each model using PM_2.5_ data with Equation (1).

**Table 5.** Coefficients of determination (*R*^2^) for each model using PM_2.5_ data with Equation (1).

| **Model** | **$R^2$** |
| --- | --- |
| Meteorological | 0.3069 |
| Meteorological + Weekend | 0.3166 |
| Meteorological + Weekend + City | 0.3846 |
| Meteorological + Weekend + City * Lockdown | 0.4768 |

From this analysis, it follows that the PM_2.5_ levels seem to be quite dependent on meteorological factors while the inclusion of city-level and lockdown effects is fundamental to improve *R*^2^ values.

### 4.3. Marginal city-specific effect for the PM_2.5_ pollutant

The minor lockdown led to overall increases in PM_2.5_ levels in the cities considered (see results of Table 4). Nevertheless, the full analysis of the proposed model requires the consideration of the city-specific coefficients omitted in Table 4. In fact, the global city-specific effects and the global period (*no lockdown*, *minor*, or *major lockdown*) effects were employed for the estimation of the city-specific marginal effects of the PM_2.5_ levels. The city-specific effects of the city-period interactions were also considered in this analysis. These results show the PM_2.5_ variations due to the COVID-19 lockdown in each city. Figure 2 shows these city-specific marginal effects with the 95% confidence intervals estimated with the Equation (1) for the PM_2.5_ pollution levels during the minor lockdown, major lockdown, and no lockdown.Madrid

**Figure 2.**
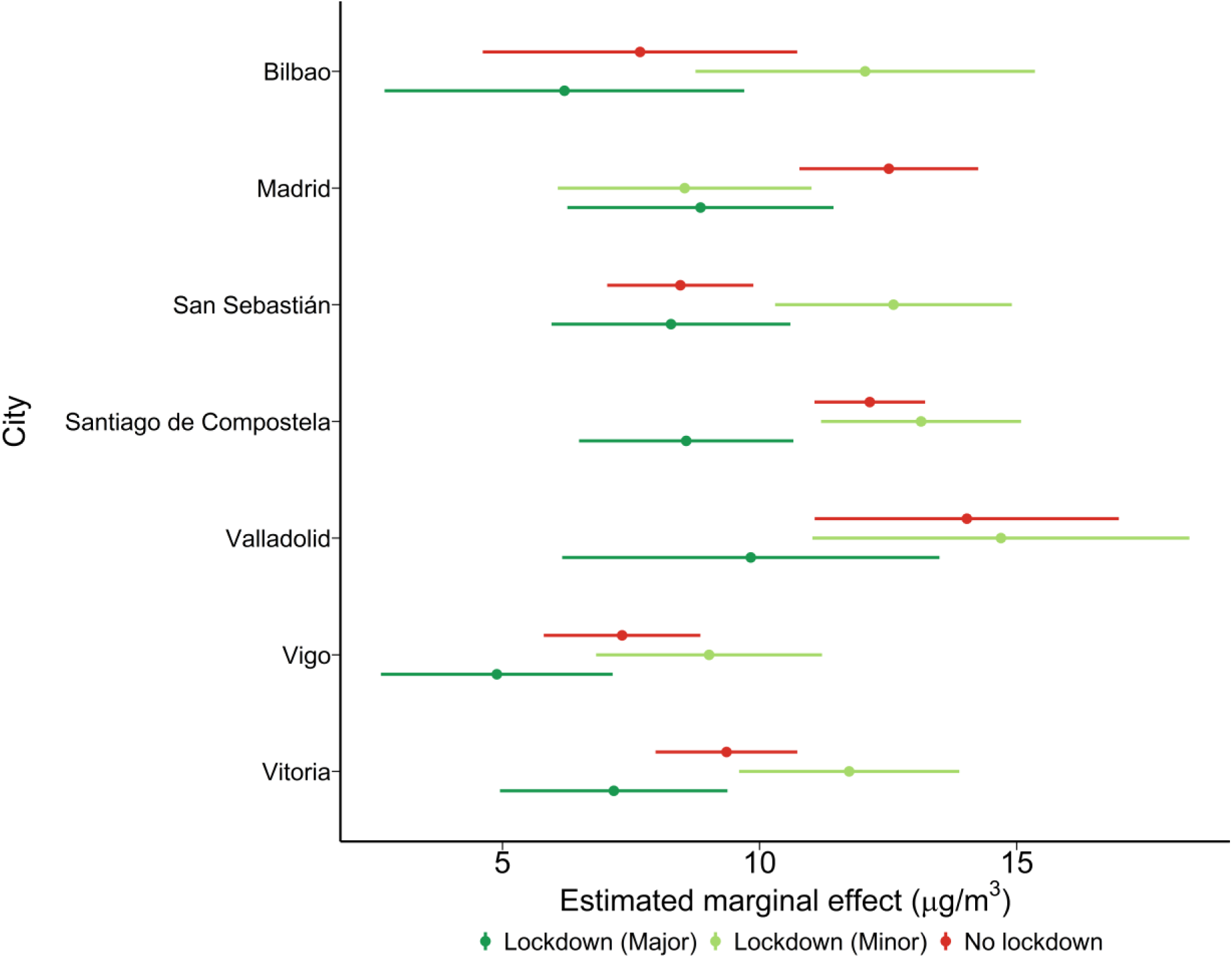
City-specific marginal effects estimated for the PM_2.5_ pollution levels during the minor lockdown, major lockdown, and no lockdown periods. Marginal effects are statistically different when there is no overlapping of the confidence intervals.

The point estimates represented in this figure show the adjusted effect of the combination of city and period under study on PM_2.5_ pollution levels. The differences between PM_2.5_ concentrations during the normal period, minor, and major lockdown were not significantly different in many of the cities. However, there was a rising trend in the minor lockdown PM_2.5_ levels in some cities considering meteorological and week-day effects, especially in Bilbao and San Sebastián. Considering that relative humidity was not included in the linear regression, these results may simply be an artifact due to high relative humidity episodes that can improve the aqueous-phase oxidation of pollutants such as SO_2_ and PM_2.5_ levels(P.Wang et al., 2020). Both are coastal cities in the north of Spain, where higher humidity is usual and are more likely to receive atmospheric particles from shipping and sea salt, two of the main sources of the pollutant analyzed. On the other hand, the major lockdown period led to a general reduction in fine particulate matter, although only one statistically significant reduction was found in Santiago de Compostela, from 12.14±0.55 to 8.57±1.06 μg/m^3^, in terms of marginal effects. This reduction of PM_2.5_ particles must be related to the drastically reduced human activity during the Spanish lockdown (P. Wang et al., 2020). However, the relationship between traffic and anthropogenic activity with particulate matter sources and particle size is not necessarily unequivocal (Nicolás et al., 2020), so that the variations shown may not always be as clear as might be expected. Nevertheless, the reduction of this fine particulate matter was not expected to be very high because these inhalable particles are emitted in large volumes and persist in the air for longer periods, which means they can spread easier(Xiao et al., 2018). In fact, these low reductions are in good agreement with our previous study of variations in air quality in terms of PM_10_ levels during this period (Briz-Redón et al., 2020).

### 4.4. Association analysis of PM_2.5_ pollution and weather patterns with COVID-19 spread

Figure 3 shows that the strict national lockdown imposed in Spain flattened the epidemic curve as expected by the Spanish authorities.Madrid Santiago de Compostela Vigo

**Figure 3.**
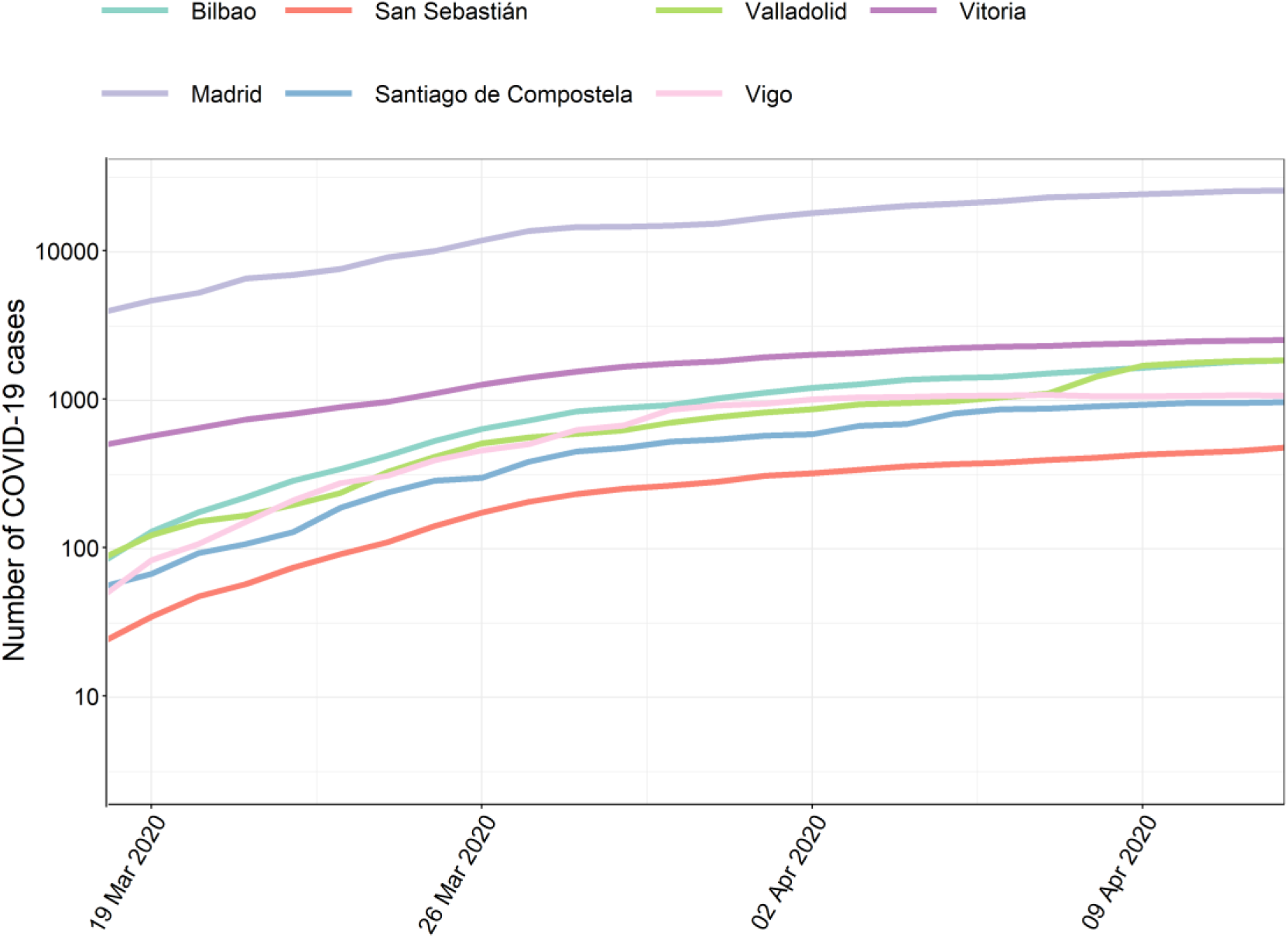
COVID-19 data used for the association study with COVID-19 accumulated cases for the seven cities considered in the study.

The relationship between PM_2.5_ exposure or meteorological factors (data are shown in Figures S1-S6 in the Supplementary material) and COVID-19 data (Figure 3) during the Spanish lockdown is shown in Figure 4.

**Figure 4.**
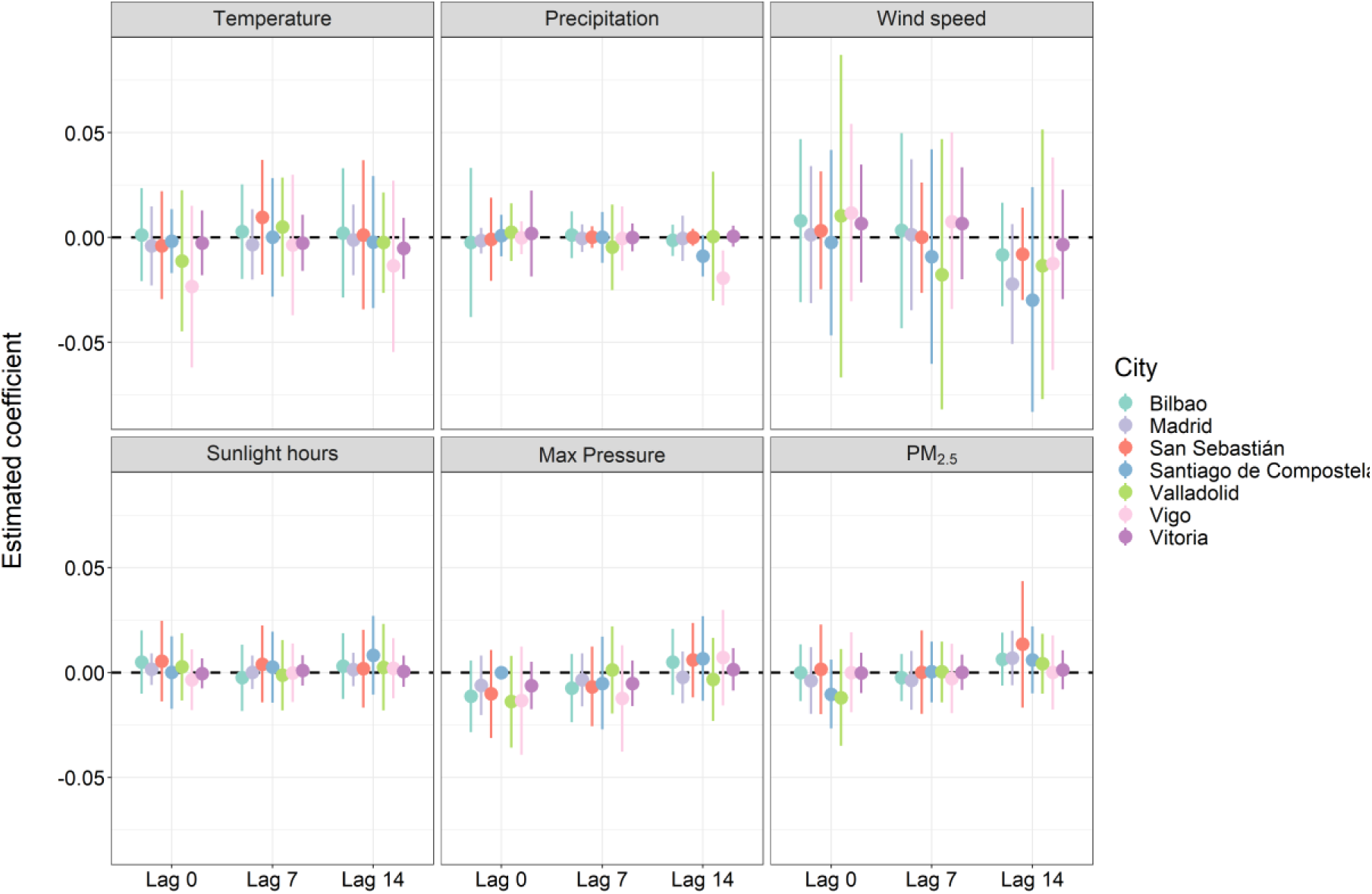
Effects of each environmental covariate and PM_2.5_ exposure on COVID-19 spread estimated for each city under study. Each dot represents the estimation of the coefficient (*β* parameter in Equation (2)), whereas the segment corresponds to the 95% credible interval associated with the estimation.

Thus, no association is observed between COVID-19 accumulated cases and PM_2.5_ levels or climate patterns, contrary to what we could expect according to recent results on the effect of particulate matter levels(Cole et al., 2020; Zhu et al., 2020; Zoran et al., 2020) or climate conditions (Chiyomaru and Takemoto, 2020; Sobur et al., 2020; Tobías and Molina, 2020). Anyhow, it is still unclear the relationship between COVID-19 transmission and PM_2.5_ exposure which is in need of further investigation (Hendryx and Luo, 2020) and many controversial results have been reported about the effect of climate on the COVID-19 spread, partly as a consequence of the different statistical methodologies chosen(Briz-Redón and Serrano-Aroca, 2020b). We have used univariate Poisson models accounting for the presence of temporal autocorrelation in the data. The inclusion of such a temporal term allows capturing the epidemic growth and reduces the chances of obtaining spurious or artefactual associations, which can easily arise when correlation coefficients are computed. The fact that many published studies are strongly based on correlation coefficients, together with the existing uncertainty about COVID-19 data, are sufficient reasons for still being cautious about the association between COVID-19 and the environment or PM_2.5_ pollution. Furthermore, the use of multivariate models that allow accounting for several of the factors (environmental or non-environmental) that are possibly implicated in COVID-19 is highly advisable, although we discarded the use of these models in this case due to the length of the time series available.

## 5. Conclusions

Air pollution was analyzed at the city-level through a regression model to determine the changes of PM_2.5_ during the Spanish lockdown while accounting for the effect of some meteorological factors. While the 4-week national lockdown reduced the COVID-19 accumulated cases in Spain, the results of this study show a significant decrease in PM_2.5_ pollution in only one city. Furthermore, no relationship between COVID-19 spread and PM_2.5_ exposure or weather patterns was found during this period in Spain.

### Supplementary material

The data of temperature (Figure S1), precipitation (Figure S2), wind velocity (Figure S3), sunlight time (Figure S4), the maximum pressure (Figure S5) and PM_2.5_ (Figure S6) levels used for the association study with COVID-19 accumulated cases for the seven cities considered in the study are shown in the Supplementary material.

## Data Availability

The data referred to in the manuscript is available under request

## Acknowledgments

The authors would like to acknowledge the University of Valencia for the *Talent attraction VLC-CAMPUS* PhD fellowship (conceded to C.B-S) and the *Fundación UCV San Vicente Mártir* for the economic support through Grants 2019-231-003UCV and 2020-231-001UCV (conceded to Á.S-A).

